# Excess mortality associated with the COVID-19 pandemic (2020-2021) in an urban community of Bangladesh

**DOI:** 10.1101/2022.11.19.22281995

**Authors:** Mohammad Sorowar Hossain, Jahidur Rahman Khan, SM Abdullah Al Mamun, Mohammad Tariqul Islam, Enayetur Raheem

## Abstract

Measurement of COVID-19-attributed mortality is vital for public health policy decisions. Unlike high-income countries, the magnitude of COVID-19-related mortality is largely unknown in many low- and middle-income countries due to inadequate COVID-19 testing capacity and a lack of robust civil registration and vital statistics systems. COVID-19-associated excess mortality was investigated in an urban setting in Bangladesh using a cemetery-based death registration dataset. A total of 6,271 deaths (3,790 male and 2,481 female) recorded between January 2015 and December 2021 were analyzed by using the Bayesian structural time series model (BSTS). During the pre-COVID-19 period, the average monthly number of deaths was 69, whereas, during the COVID-19 period, this number significantly increased to 92. Overall, according to model-based results, during COVID-19 period, the number of deaths increased on average by 17% (95% CrI: -18%, 57%): males 29% (95 % CrI: -15%, 75%) and 2.9% for females (95% CrI: -61%, 70%). This first-of-its-kind study in Bangladesh has revealed the excess mortality due to the COVID-19 pandemic (2020-2021) in an urban community. It appears that cemetery-based death registration could help track various crises (e.g., COVID-19), especially when collecting data on the ground is challenging for resource-limited countries.

## Background

Understanding the magnitude of COVID-19-attributed mortality is crucial for public health policy decisions to reduce mortality and prevent future crises. Unlike high-income countries, underreporting deaths is a genuine concern for most low- and middle-income countries (LMICs) due to inadequate COVID-19 testing capacity and a lack of robust civil registration and vital statistics systems[1]. Consequently, the extent of COVID-19-related mortality is unclear in many LMICs, including Bangladesh. Excess mortality is a commonly used indicator of the observed death from any cause in comparison to that anticipated based on historical averages. Using a cemetery-based death registration dataset, we sought to quantify the magnitude of excess mortality in an urban community in Bangladesh.

## Methods

### Study setting

COVID-19-associated excess mortality was analyzed for Jamalpur town (also known as pauroshova, one of the 64 district towns in Bangladesh), an urban setting which is located 142 kilometers northwest of the capital city, Dhaka. In the 53.3 km sq study area, more than 180,358 inhabitants (population density ∼ 3385/km sq) with an annual growth rate of 1.7% live here based on 2015 consensus data[2]. Notably, Bangladesh has no national system for registering deaths and determining their causes. Most deaths occur at home and therefore the cause of death is primarily unknown. The death certificate issued by the public hospitals remains questionable due to the lack of a proper medical record-keeping system[3,4]. Since 2010, death registration could be done directly by visiting a registration office (such as pauroshova). While it is mandatory to show a death certificate for property succession, claiming a pension and insurance, and nominated bank deposit, families with a lower economic background are not often interested in making death registration. Because of space constraints and the high land prices in towns, almost all families prefer public cemeteries to bury their loved ones. On the other hand, in rural areas, people generally bury the deceased in private cemeteries. Even though people migrate outside the community for livelihood, they are generally buried in the local community cemetery upon their death.

### Dataset

We extracted available information (age, sex, and month of demise) of individual death cases from the pauroshova death registry books. Jamalpur town collects self-reported information on death cases registered at the existing four Muslim cemeteries and one Hindu cremation place (supplement). The dataset contained 10,482 deaths (January 2011–December 2021); 6 incomplete records were removed; 1,610 records were excluded from analysis whose age was under 35 years (since over 96% COVID-19 related official deaths occurred at >35 years)[5], resulting in 8,866 deaths. Finally, 6,271 deaths (3,790 male and 2,481 female) recorded between January 2015 and December 2021 were analyzed.. The fatalities were measured as the monthly count of deaths for any cause (outcome measure). The exposure period (COVID-19) was defined as March 2020 to December 2021 (COVID-19 period), and the pre-exposure period (pre-COVID-19 period) was defined as January 2015 to February 2020.

### Statistical analysis

To measure the impact of COVID-19 on mortality, the Bayesian structural time series model was used in this study, which has been described in detail elsewhere[6]. It is a stochastic state-space model that can incorporate trend, seasonality, and regression components[7]. Using the R, the point effects (i.e., relative effects = [∑predicted-∑observed]/∑predicted) and their 95% credible intervals (CrIs) were generated by comparing estimated forecasts to observed trends across 40,000 Markov Chain Monte Carlo iterations (10% burn-in period) to measure the effects of COVID-19. This study was approved by the Institutional Review Board at Biomedical Research Foundation, Bangladesh (BRF/ERB/2020/E03).

## Results

**Table 1** compares the monthly average number of fatalities before and during the COVID-19 outbreak, overall and individually for male and female. During the pre-COVID-19 period, the average monthly number of deaths was 69, whereas during the COVID-19 period, this figure increased by 23 and reached 92. During the COVID-19 period, the average monthly number of male and female deaths increased significantly by 16 (from 40.9% to 57.0%) and 7 (from 27.6% to 35.0%), respectively. This suggests that males experienced a greater average monthly death than women.

**Table 1.** The difference in the monthly average number of deaths between pre-COVID-19 and COVID periods, as well as a model-based estimate of the impact of COVID-19.

| **Number of deaths** | **Pre-COVID**<br>**(Jan 2015 to Feb 2020)** | **During COVID**<br>**(Mar 2020 to Dec 2021)** | **Difference<sup>1</sup>** | **Relative effect**<br>**95% Credible Interval<sup>2</sup>** |
| --- | --- | --- | --- | --- |
| Total | 68.5 | 91.9 | 23.4* | 17% (-18%, 57%) |
| Male | 40.9 | 57.0 | 16.1* | 29% (-15%, 75%) |
| Female | 27.6 | 35.0 | 7.4* | 2.9% (-61%, 70%) |

<sup>1</sup>Difference between the monthly average deaths, <sup>2</sup>BSTS model based results on the impact of COVID-19
\*p<0.001, based on Wilcoxon rank-sum test

Figure 1 shows the differences between observed and model predicted deaths (dotted line). There was a notable rise in month deaths during the COVID-19 period, specific months (e.g., August-September 2020). The cumulative difference between observed and predicted deaths in the COVID-19 period shown in Figure 2 indicated an increase in the number of deaths, particularly among males. The summary model-based results for the full and stratified by gender analyses are shown in Table 1. Overall, according to model-based results, during COVID-19 period, the number of deaths increased on average by 17% (95% CrI: -18%, 57%): males 29% (95 % CrI: - 15%, 75%) and 2.9% for females (95% CrI: -61%, 70%). The results suggest that the commencement of the COVID-19 pandemic related to an increase in the number of deaths in the studied location. Even though the results were not statistically significant, the considerable increase in deaths during the COVID-19 period may be indicative of an excess of mortality.

**Figure 1:**
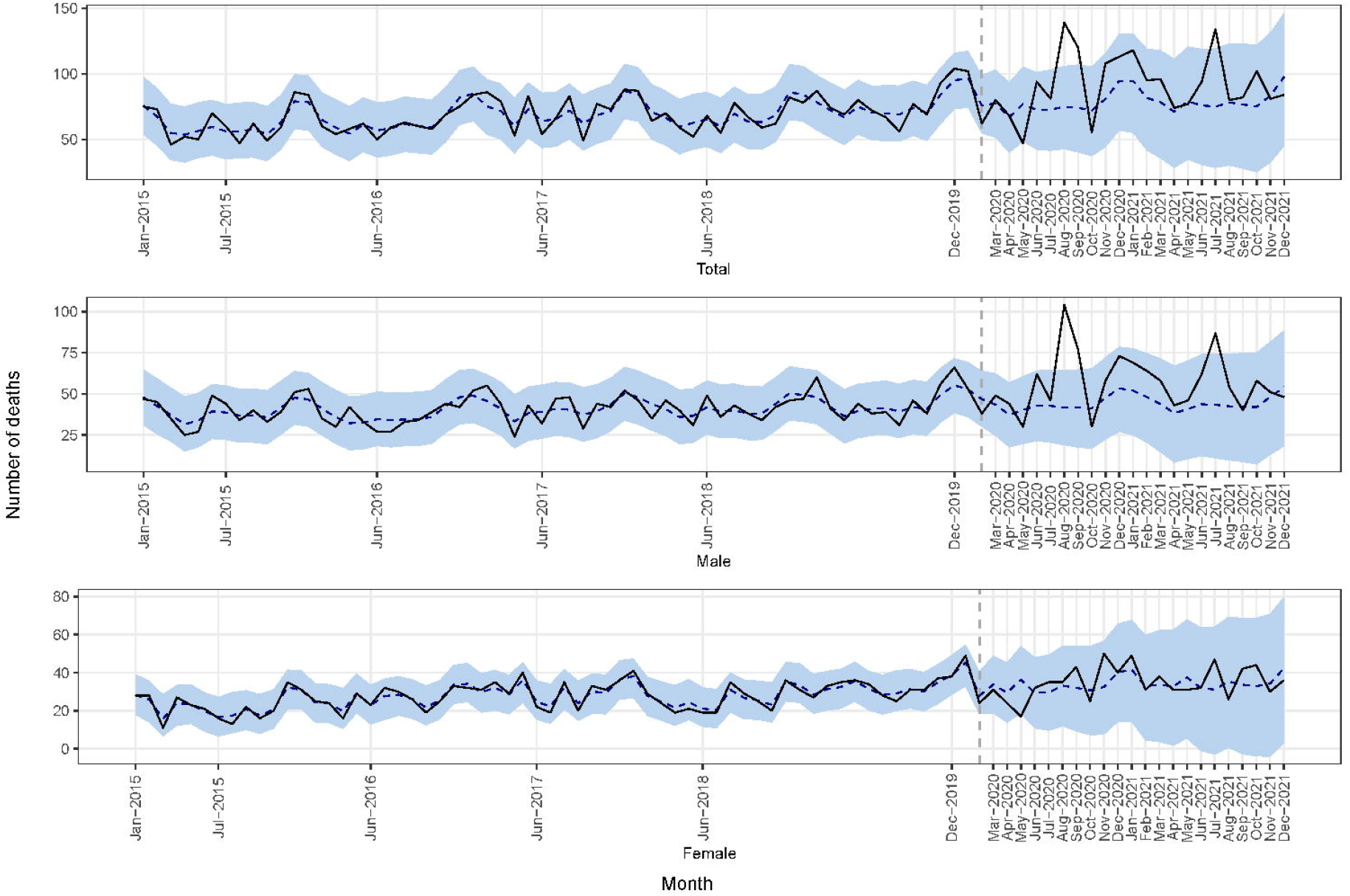
The monthly number of observed and predicted deaths (total, male, and female) observed (solid line) and predicted (dotted line) time series and 95% credible intervals (blue areas) according to the Bayesian structural time series.

**Figure 2:**
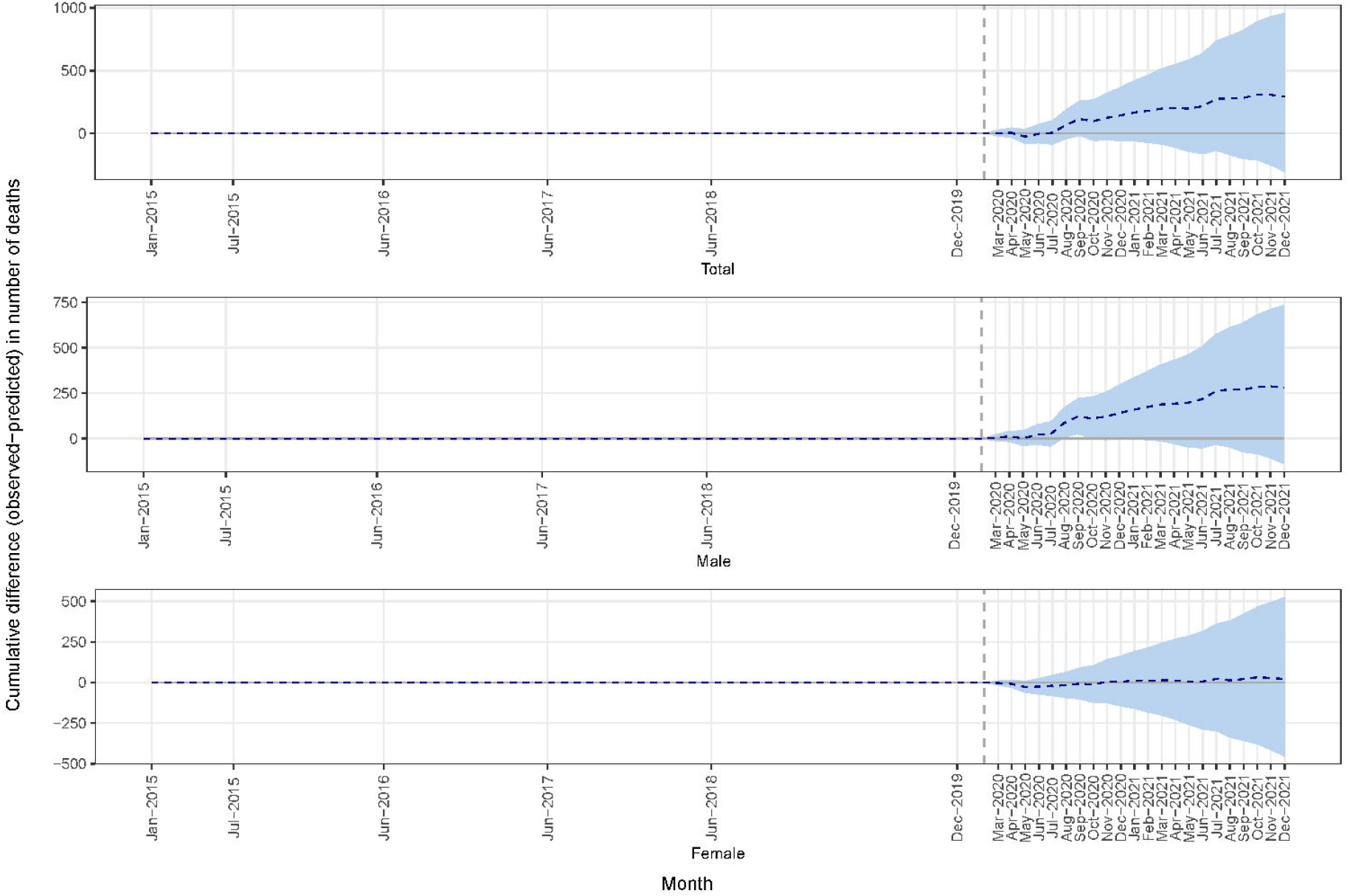
The cumulative difference between observed and predicted deaths (dotted line) in the COVID period

## Discussion

This first-of-its-kind study in Bangladesh has revealed the excess mortality in an urban community due to the COVID-19 pandemic over a period between 2020 and 2021. Based on household surveys, two prior studies so far published from Bangladesh focused on rural settings. In one study, all-cause mortality was lower during the early months of the COVID-19 pandemic (2020) as compared to 2019[8], while no excess mortality was reported in another study, except higher mortality reported among 65 years or older during the first year of the pandemic [9]. Consistent with national sex-specific COVID-19 deaths in Bangladesh, this study also found excess mortality among males[5]. Novel model-based results on excess mortality were not statistically different from the null for the entire COVID-19 period, but individual months or shorter periods had a significant impact. Random fluctuations of observed series, a lengthy COVID-19 period, and the absence of sufficient control variables may have contributed to the overall significance of the findings. Our findings are not generalizable to all urban settings in the country. However, important demographic and socio-economic factors of Jamalpur are comparable to other 33 district towns (out of 64) in Bangladesh[10]. It appears that cemetery-based death registration could help track various crises (e.g., COVID-19), especially when collecting data on the ground is challenging for resource-limited countries.

## Data Availability

All data produced in the present work are contained in the manuscript

## Declaration

### Authors’ contributions

MSH conceived the idea. MSH, JRK, ER designed the study. All co-authors contributed data acquisition, analysis, and interpretation and provided critical feedback

### Conflict of interest statements

We declare that there is no conflict of interest

### Role of funding source

No funding was associated with this work

### Ethics committee approval

This study was approved by the Institutional Review Board at Biomedical Research Foundation, Bangladesh (BRF/ERB/2020/E03).

## Notes

### Competing Interest Statement

The authors have declared no competing interest.

### Funding Statement

No funding was associated with this project

